# Intravenous Immunoglobulin (IVIG) Significantly Reduces Respiratory Morbidity in COVID-19 Pneumonia: A Prospective Randomized Trial

**DOI:** 10.1101/2020.07.20.20157891

**Authors:** George Sakoulas, Matthew Geriak, Ravina Kullar, Kristina L. Greenwood, MacKenzie Habib, Anuja Vyas, Mitra Ghafourian, Venkata Naga Kiran Dintyala, Fadi Haddad

**Author notes:** Corresponding author: George Sakoulas, MD, UCSD School of Medicine Biomedical Research Facility II, Room 4114, 9500 Gilman Drive, Mail Code 0760, La Jolla, CA 92093-0760. **Conflicts of Interest:** None. **Funding:** IVIG (Octagam 10%) was provided by Octapharma USA, Hoboken, NJ.

## Abstract

**Background:** Interventions mitigating progression to mechanical ventilation in COVID-19 would markedly improve outcome and reduce healthcare utilization. We hypothesized that immunomodulation with IVIG would improve oxygenation and reduce length of hospital stay and progression to mechanical ventilation in COVID-19 pneumonia.

**Methods:** Patients with COVID-19 were randomized 1:1 to prospectively receive standard of care (SOC) plus IVIG 0.5 g/kg/day × 3 days with methylprednisolone 40 mg 30 minutes before infusion versus SOC alone.

**Results:** 16 subjects received IVIG plus SOC and 17 SOC alone. The median age was 51 years for SOC and 58 years for IVIG. APACHE II scores and Charlson comorbidity indices were similar for IVIG and SOC (median 7.5 vs 7 and 2 for both, respectively). Seven SOC versus 2 IVIG subjects required mechanical ventilation (p=0.12, Fisher exact test). Among subjects with A-a gradient of >200 mm Hg at enrollment, the IVIG group showed i) a lower rate of progression to requiring mechanical ventilation (2/14 vs 7/12, p=0.038 Fisher exact test), ii) shorter median hospital length of stay (11 vs 19 days, p=0.01 Mann Whitney U), iii) shorter median ICU stay (2.5 vs 12.5 days, p=0.006 Mann Whitey U), and iv) greater improvement in PaO_2_/FiO_2_ at 7 days (median [range] change from time of enrollment +131 [+35 to +330] vs +44·5 [-115 to +157], p=0.01, Mann Whitney-U test) than SOC.

**Conclusion:** This pilot prospective randomized study comprising largely of Latino patients showed that IVIG significantly improved hypoxia and reduced hospital length of stay and progression to mechanical ventilation in COVID-19 patients with A-a gradient >200 mm Hg.

## Introduction

COVID-19 infection, as is common with many types of viral and atypical infections, is characterized by a biphasic illness of a relatively mild protean phase driven by viral replication resolving in about a week, and second phase, driven by the immune response. The second phase may lead to potentially catastrophic disease manifestations requiring hospitalization and high-level medical care characterized by acute respiratory distress syndrome (ARDS), vasculitis with thrombotic complications, and multi-organ involvement.^1-4^ It is not surprising, therefore, that the clinical effects of remdesivir, and hydroxychloroquine, molecules with defined anti-viral activities in vitro and in vivo, are modest at best in treating COVID-19 hospitalized patients.^5-8^ Therefore, from a pathophysiological standpoint, clinically meaningful therapies for COVID-19 will likely emerge from immunomodulation. Several studies are under way investigating immunosuppressive agents, but many of these agents focus on potent inhibition of a single immunological target, including the interleukin-6 receptor inhibitor, tocilizumab.^9,-14^ Unfortunately, many complex biological systems including the human immune response to infection, are highly redundant and therefore, attenuation of a single target may be bypassed, compromising clinical effectiveness of an agent which may be promising in laboratory settings.^15^ Furthermore, potent inhibition of specific pathways may come at a price of targeted immunosuppression and an increased risk of opportunistic infection, as seen in patients treated with these agents for autoimmune disease.^16^

IVIG has been found to have broad therapeutic applications for the treatment of a variety of inflammatory, infectious, autoimmune, and viral diseases including Kawasaki disease. IVIG may modulate the immune response via multiple mechanisms including blocking a wide array of pro-inflammatory cytokines that potentially lead to severe inflammatory responses as well as Fc-gamma receptor binding of activated macrophages.^18^ There are published reports retrospectively showing potential benefit of IVIG treatment for COVID-19 ARDS in adults and the associated Kawasaki-like illness in children.^19-24^ This is the first study to prospectively evaluate the addition of IVIG to otherwise standard treatment for adults with moderate to severe hypoxemia secondary to COVID-19.

## Methods

### Study design

This was an open-label randomized controlled trial performed at two hospital centers: Sharp Memorial Hospital (San Diego, CA) and Sharp Grossmont Hospital (La Mesa, CA). The research protocol was approved by the Internal Review Board of the participating hospitals prior to patient enrollment and was registered on clinicaltrials.gov (April 28, 2020;; NCT04411667). All participants provided informed consent electronically.

### Study population

Adult patients ≥ 18 years of age presenting with COVID-19 infection confirmed by positive polymerase chain reaction testing for SARS-CoV2 genome in nasopharyngeal or oropharyngeal swab sample were considered for inclusion if they demonstrated moderate to severe hypoxia (sPO_2_ ≤96% on ≥ 4 liters O_2_ by nasal cannula) but not on mechanical ventilation. This corresponds to FiO_2_ of 37% to maintain a PaO2 of 90 mm Hg (alveolar-arterial [A-a] gradient of 120 mm Hg or PaO_2_/FiO_2_ 243).

### Randomization and Treatments

After informed consent was obtained, subjects were randomized 1:1 into treatment arm or standard of care (SOC) control arm. SOC consisted of the subject remaining on or being eligible for any treatment not part of a randomized clinical trial at the time of enrollment. On May 13, 2020 and afterwards, this included the use of remdesivir. Subjects were also allowed to receive convalescent plasma therapy as part of the nationally available compassionate use registry. The IVIG treatment arm consisted of the subject receiving IVIG (Octagam 10% provided by Octapharma USA, Inc) 0.5 g/kg daily for 3 days beginning on the day of enrollment in addition to SOC. For subjects not already receiving glucocorticoid therapy, enrolled treatment arm subjects received methylprednisolone 40 mg IV once 30 minutes before IVIG to mitigate headache commonly experienced after IVIG therapy. Enrollment in other clinical trials and the use of off-label agents (eg. tocilizumab) was not allowed while the subject was enrolled and monitored for progression to the endpoint of i) respiratory failure requiring receipt of mechanical ventilation (a composite of either receiving ventilation or the subject status changed to a do not resuscitate/do not intubate resulting in progressive respiratory failure and death) or ii) death from non-respiratory causes prior to receipt of mechanical ventilation. If the subject progressed to mechanical ventilation, receipt of off-label agents and/or enrollment in other clinical trials was allowed. Subject hospital course was followed until hospital discharge or for 30-days in the hospital after enrollment, whichever came first, for the purpose of total and intensive care unit (ICU) days of hospital days.

### Clinical data extraction and analysis

Relevant clinical and laboratory information was captured to allow for group comparisons, including the calculation of Charlson comorbidity index (https://www.mdcalc.com/charlson-comorbidity-index-cci) and APACHE II acute illness severity score (https://www.mdcalc.com/apache-ii-score#next-steps). The alveolar-arterial (A-a) gradient was calculated (https://www.mdcalc.com/a-a-o2-gradient) for each subject at the time of enrollment based on arterial blood gases when available or based on PaO_2_ extrapolated SpO_2_ and fraction of inspired oxygen (FiO_2_). Interleukin-6 was measured from blood 24-48 hours after enrollment (https://ltd.aruplab.com/Tests/Pub/0051537).

### Statistical analysis

The initial goal of the pilot study was to enroll 20 patients (10 per arm). After 20 patients had reached the composite endpoint or discharged from the hospital, the data was reviewed by a data safety monitoring board (DSMB) consisting of 2 hospitalist physicians, 1 critical care physician, and 1 pharmacist/statistician. The DSMB voted to continue the study until the Phase 3 randomized placebo-controlled multi-center study of IVIG in COVID-19 (Octagam 10% therapy in COVID-19 patients with severe disease;; clinicaltrials.gov, NCT04400058, May 22, 2020) became available. All analyses were performed on the intent to treat population. Statistical differences in rates of receipt of mechanical ventilation and other categorical or ordinal variables were calculated using Fisher exact test, and differences in continuous variables were calculated using Mann Whitney-U.

## Results

### Baseline patient demographics and clinical characteristics

Between May 1 and June 16, 2020, 34 patients were randomized into the study as shown in Figure 1 (17 SOC, 17 IVIG). Immediately after randomization and notification of the principal investigator, one subject was immediately deemed unevaluable by the principal investigator and excluded due to a high risk of bacterial superinfection (elevated absolute neutrophil count of 9900/mm^3^ and concomitant procalcitonin of 1.45 ng/mL).^1,25^ The characteristics and demographics of the 17 SOC and 16 IVIG subjects are shown in Table 1. The subjects were well-matched with respect to age and underlying comorbidities. Consistent with the pattern of COVID-19 infection in San Diego, >80% of enrolled subjects in each arm were Latino and predominantly male. Concomitant therapies were also evenly distributed in both arms, with about half of the enrolled subjects in each arm receiving remdesivir. Convalescent plasma was administered to 5 subjects (3 SOC, 2 IVIG). Glucocorticoid therapy was part of the IVIG protocol and used as a premedication (40 mg IV methylprednisolone before each IVIG dose for 3 days) whereas 10 of 17 SOC subjects received some glucocorticoid therapy. Five IVIG subjects received additional glucocorticoid therapy beyond the methylprednisolone per protocol dosing. The SOC subjects receiving glucocorticoids received them for a median of 11 days.

**Table 1.**
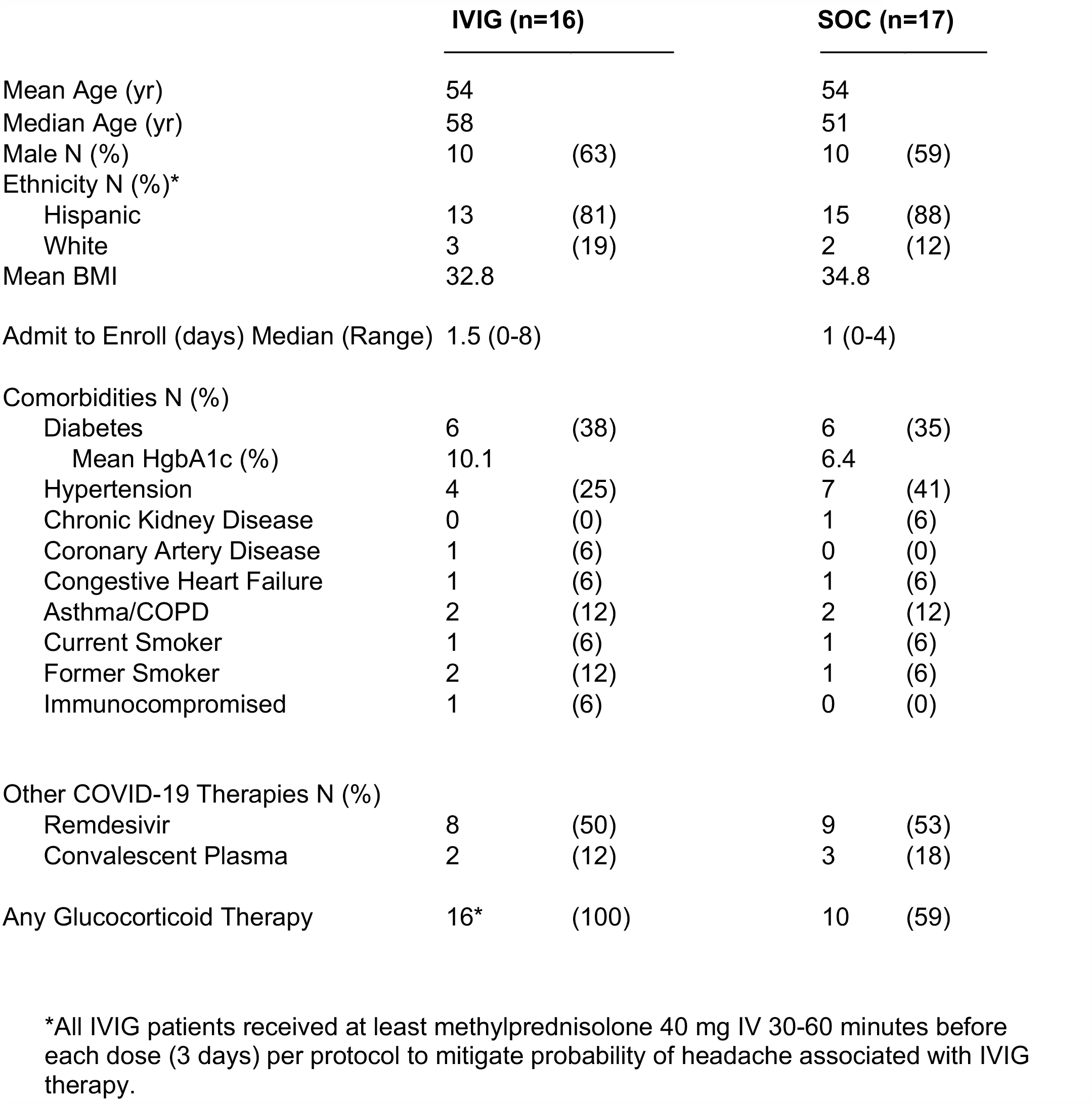
Enrolled patient demographics and characteristics.

|  | IVIG (n=16) |  | SOC (n=17) |  |
| --- | --- | --- | --- | --- |
| Mean Age (yr) | 54 |  | 54 |  |
| Median Age (yr) | 58 |  | 51 |  |
| Male N (%) | 10 | (63) | 10 | (59) |
| Ethnicity N (%)* |  |  |  |  |
| Hispanic | 13 | (81) | 15 | (88) |
| White | 3 | (19) | 2 | (12) |
| Mean BMI | 32.8 |  | 34.8 |  |
| Admit to Enroll (days) Median (Range) | 1.5 | (0-8) | 1 | (0-4) |
| Comorbidities N (%) |  |  |  |  |
| Diabetes | 6 | (38) | 6 | (35) |
| Mean HgbA1c (%) | 10.1 |  | 6.4 |  |
| Hypertension | 4 | (25) | 7 | (41) |
| Chronic Kidney Disease | 0 | (0) | 1 | (6) |
| Coronary Artery Disease | 1 | (6) | 0 | (0) |
| Congestive Heart Failure | 1 | (6) | 1 | (6) |
| Asthma/COPD | 2 | (12) | 2 | (12) |
| Current Smoker | 1 | (6) | 1 | (6) |
| Former Smoker | 2 | (12) | 1 | (6) |
| Immunocompromised | 1 | (6) | 0 | (0) |
| Other COVID-19 Therapies N (%) |  |  |  |  |
| Remdesivir | 8 | (50) | 9 | (53) |
| Convalescent Plasma | 2 | (12) | 3 | (18) |
| Any Glucocorticoid Therapy | 16* | (100) | 10 | (59) |

\*All IVIG patients received at least methylprednisolone 40 mg IV 30-60 minutes before each dose (3 days) per protocol to mitigate probability of headache associated with IVIG therapy.

**Figure 1.**
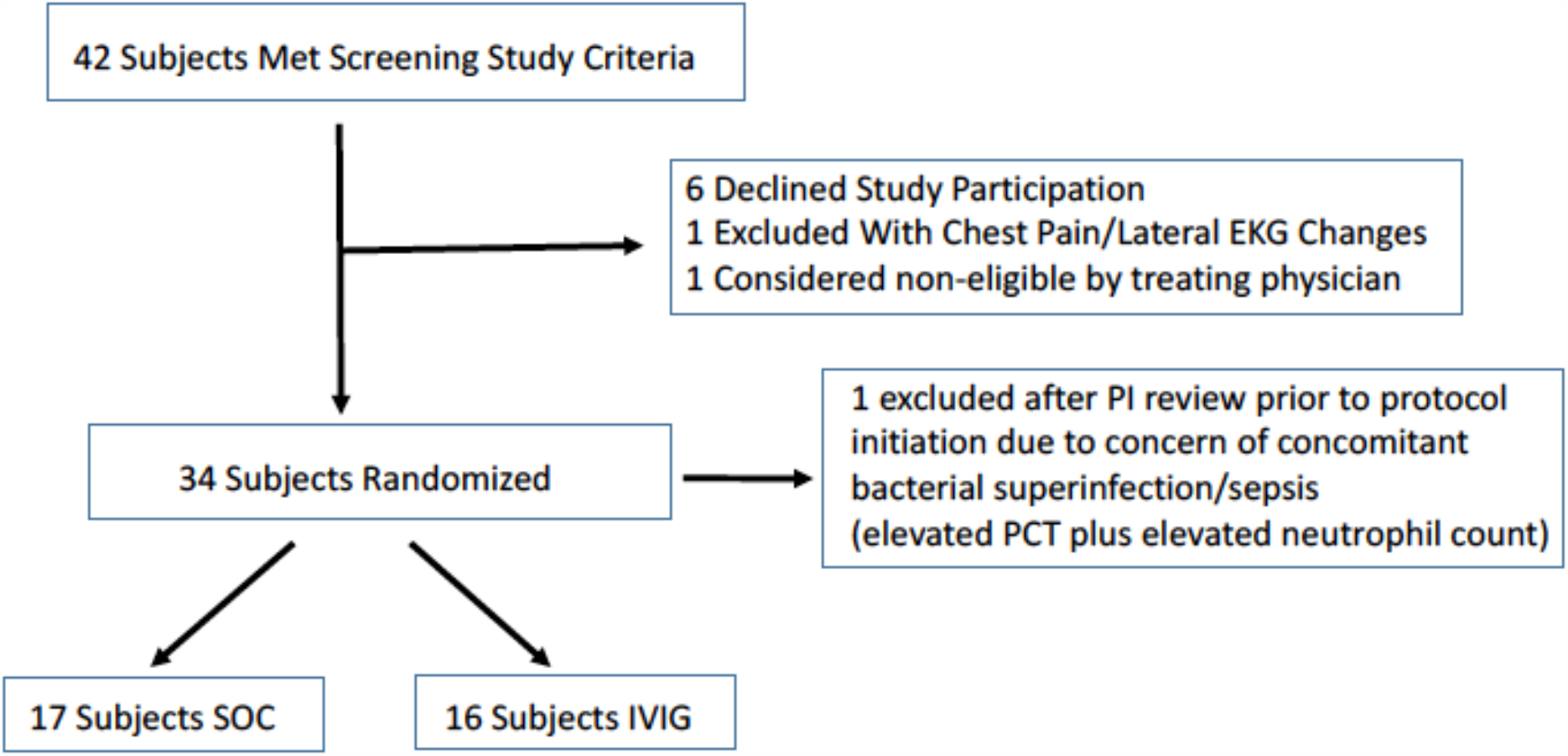
Consort Diagram of Enrolled Patients

Laboratory data was well matched between the two treatment groups, as shown in Table 2. Significant leukocytosis > 12,000/mm^3^ was uncommon, present in only 4 total subjects (3 IVIG and 1 SOC) and procalcitonin averaged 0.25 ng/mL for both groups, with a maximum of 0.80 ng/mL. D-dimer concentrations at the time of enrollment were numerically higher in the IVIG group compared to the SOC, with means of 1456 and 758 ng/mL, respectively (normal <500 ng/mL). Median (range) of D-dimer concentrations were 827 (386-7028) and 691 (283-1657) in IVIG and SOC groups, respectively. One IVIG and 4 SOC subjects had normal D-dimer concentrations at enrollment.

**Table 2.**
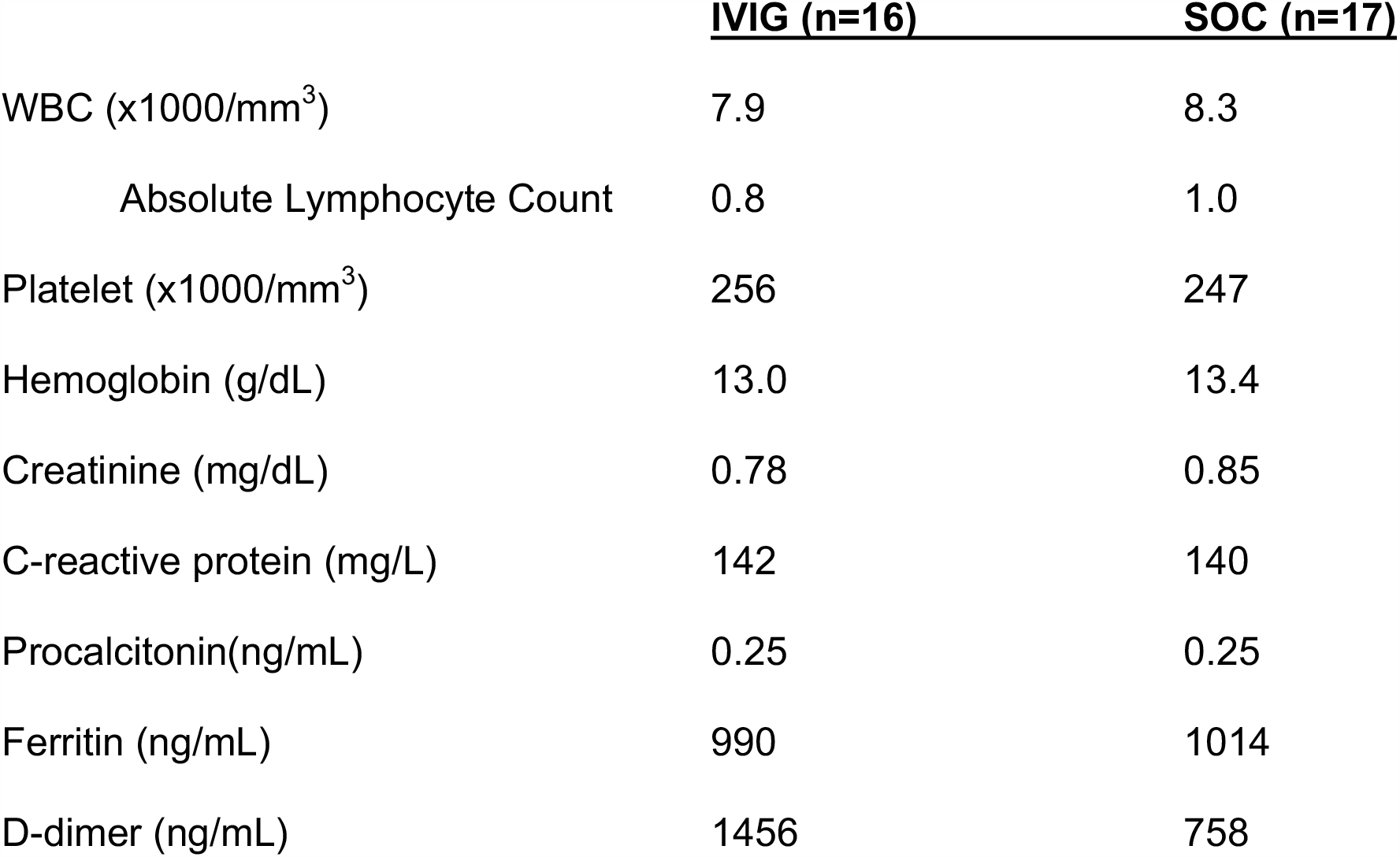
Mean Values of Relevant Laboratory Data for Study Groups.

### Clinical outcomes

The two patient groups were well-matched with respect to their Charlson comorbidity index and severity of illness APACHE 2 scores (Figure 2). Median Charlson index was 2 for both groups. Median APACHE 2 score was 7 for SOC and 7.5 the IVIG study group. Figure 2 demonstrates the subjects in the control SOC and treatment IVIG group that developed a need for mechanical ventilation after study enrollment, denoted as the red data points. An overall trend is for receipt of mechanical ventilation in the IVIG group in the 2 patients with highest comorbidity and illness severity scores, whereas in the SOC group mechanical ventilation requirement developed across the entire spectrum of scores.

**Figure 2.**
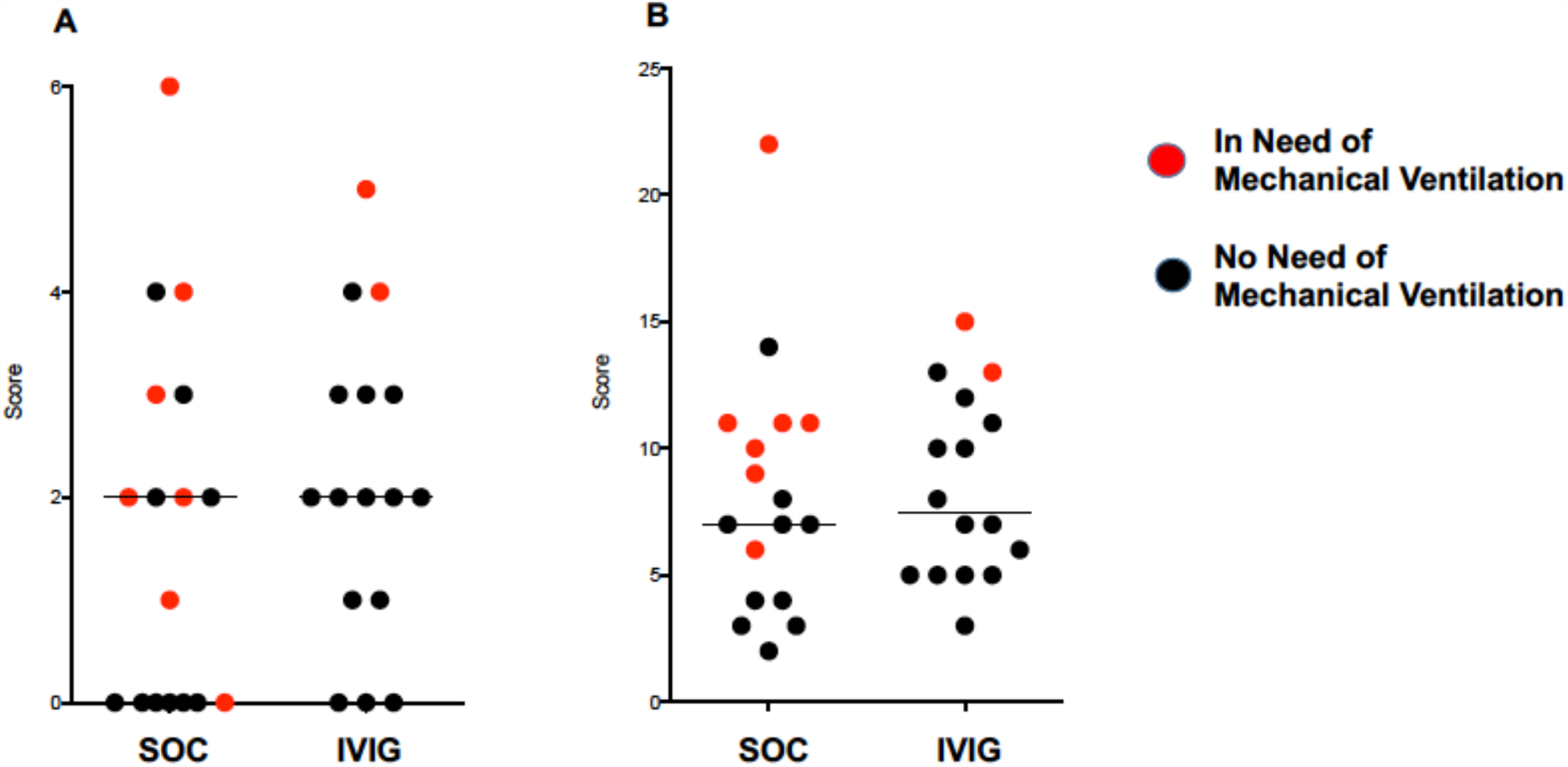
Distribution of Charlson comorbidity index (A) and APACHE 2 scores (B) of enrolled study subjects in both treatment arms, showing even distribution of chronic illness and acute severity of illness. Horizontal bars denote median values. Red points indicate patients who ultimately required a need for mechanical ventilation.

Among the entire enrolled and evaluated subjects in each arm, 2 patients in the IVIG arm and 7 patients in the control SOC arm developed a need for mechanical ventilation. Among these 7 SOC patients, 6 received ventilation and 1 was made a ‘do not intubate’ and the patient expired within 24 hours. The difference in receipt of mechanical ventilation was not statistically significant between the two groups (p=0.12, Fisher exact test). Among subjects whose respiratory failure progressed to the need for mechanical ventilation, 1 of 2 IVIG subjects and 2 of 3 control subjects also received concomitant convalescent plasma. Concomitant glucocorticoid therapy was given to 5 of the 7 control subjects who progressed to mechanical ventilation. The use of remdesivir was dependent on its availability after May 13, 2020 and therefore all patients admitted after that date whose respiratory failure progressed to needing mechanical ventilation received it. This resulted in 1 of 2 IVIG patients and 3 of 7 SOC subjects that required ventilation receiving remdesivir.

Alveolar-arterial (A-a) gradients were calculated directly from arterial blood gas or estimated based on PaO_2_ and FiO_2_ measurement. Based on an increase in APACHE II severity of illness scoring associated with achieving and A-a gradient of > 200 mm Hg (+2 points), subjects were stratified into those with A-a gradient ≤ 200 mm Hg or >200mm Hg, corresponding to a PaO_2_/FiO_2_ of <140, or the approximate requirement of 6 liters O_2_ by nasal cannula for a PaO2 of ≤92%. As shown in Figure 3A, none of the 7 subjects (5 SOC, 2 IVIG) with A-a gradient ≤ 200 mm Hg progressed to mechanical ventilation, but for subjects with A-a gradient of > 200 mm Hg at enrollment, the progression to mechanical ventilation was 7/12 (58%) in the SOC control arm vs 2/14 (14%) in the IVIG, a difference that was statistically significant (p=0.038, Fisher Exact test).

**Figure 3.**
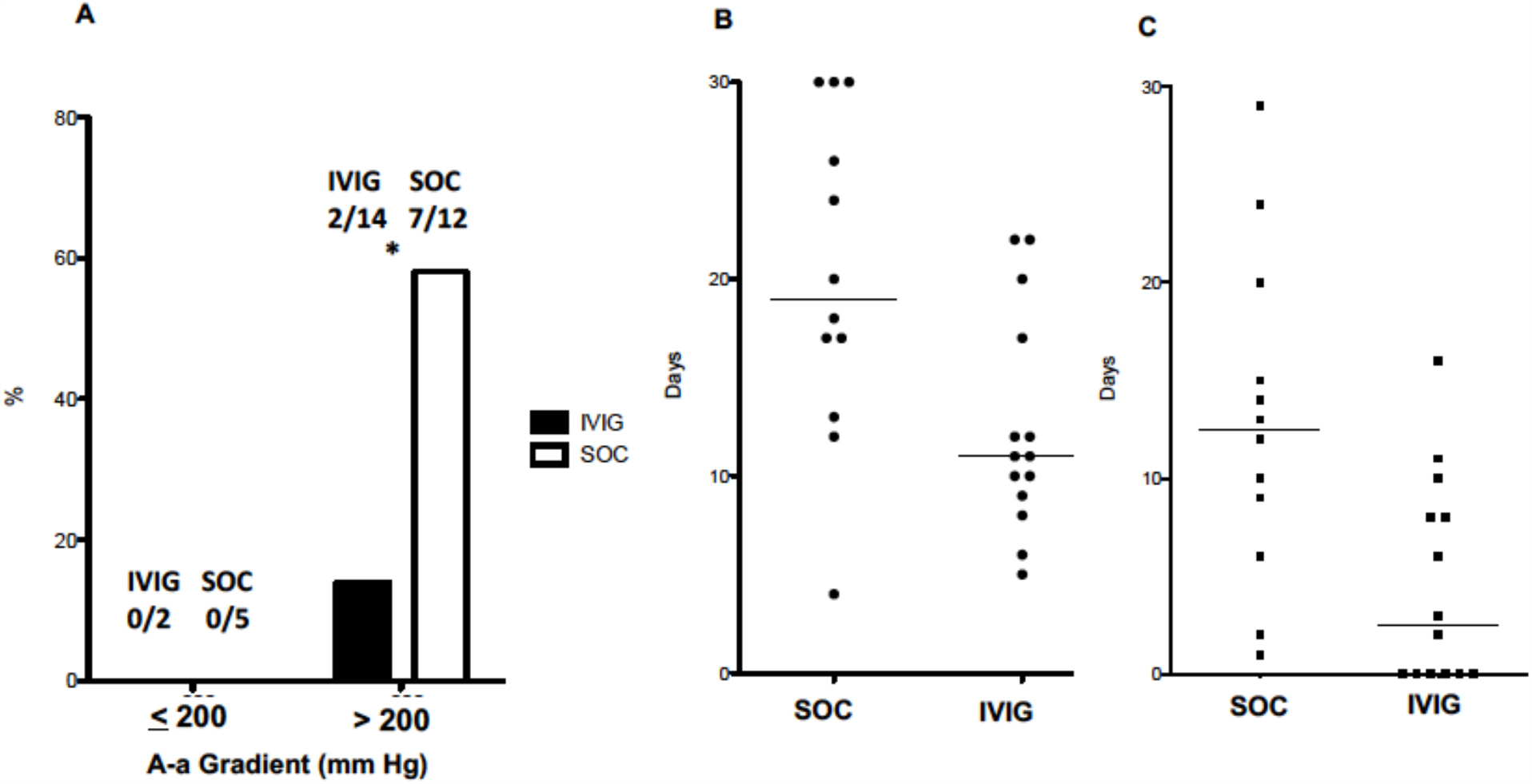
A. Rates of mechanical ventilation in study subjects stratified by A-a gradient. Among patients with A-a gradient > 200 mm Hg, receipt of IVIG reduced rates of mechanical ventilation (*p=0.038, Fisher Exact test). B. Total length of hospital stay (days) among patients in SOC vs. IVIG with A-a gradient >200 mm Hg. Median stay (horizontal bar) SOC 19 days vs. IVIG 11 days, p=0.01 Mann Whitney U test. C. Length of ICU stay (days) among patients in SOC vs. IVIG with A-a gradient >200 mm Hg. Median (horizontal bar) stay SOC 12.5 days vs. IVIG 2.5 days, p=0.006 Mann Whitney U test.

Evaluation of overall hospital course, including length of hospital stay and length of ICU stay also was highly dependent on A-a gradient stratification. Among the 7 patients with A-a gradient ≤ 200 mm Hg, no patient required ICU stay during their illness and length of hospital stay were 3-8 days. However, for the subjects with A-a gradient > 200 at enrollment, median length of hospital stay was 19 (range 4-30) and 11 (range 5-22) days for SOC and IVIG groups, respectively (p=0.013, Mann Whitney U test, Figure 3B). Median ICU stay were 12.5 days (range 1-29) and 2.5 days (range 0-16) for SOC and IVIG, respectively (p=0.006, Mann Whitney U test, Figure 3C). Total ventilator patient-days were 98 days for SOC (5.8 days/patient enrolled) and 23 days for the IVIG group (1.4 days/patient enrolled). The supplementary Figure shows schematically the hospital stays from the time of admission of the 33 enrolled patients with respect to medical floor and ICU stays with and without mechanical ventilation.

Improvement in oxygenation was evaluated by examining the PaO_2_/FiO_2_ ratio at the day of enrollment and 7 days after enrollment in individual subjects for SOC (Figure 4A) and IVIG (Figure 4B). Differences in day 7 PaO_2_/FiO_2_ minus enrollment PaO_2_/FiO_2_ are shown in Figure 4C for both groups, with negative numbers representing worsening in oxygenation. Among the entire subject population, median improvement PaO_2_/FiO_2_ for IVIG was +153 (range +35 to + 330) and was better than SOC +90 (range −115 to +280), but did not reach statistical significance (p=0·057, Mann Whitney U test). However, when refocusing on the higher-risk patients with A-a gradient >200 at enrollment, median improvement of PaO_2_/FiO_2_ for IVIG was significantly greater than SOC, with median (range) of +131 (+35 to +330) versus +44·5 (−115 to +157) (p=0·01, Mann Whitney-U test).

**Figure 4.**
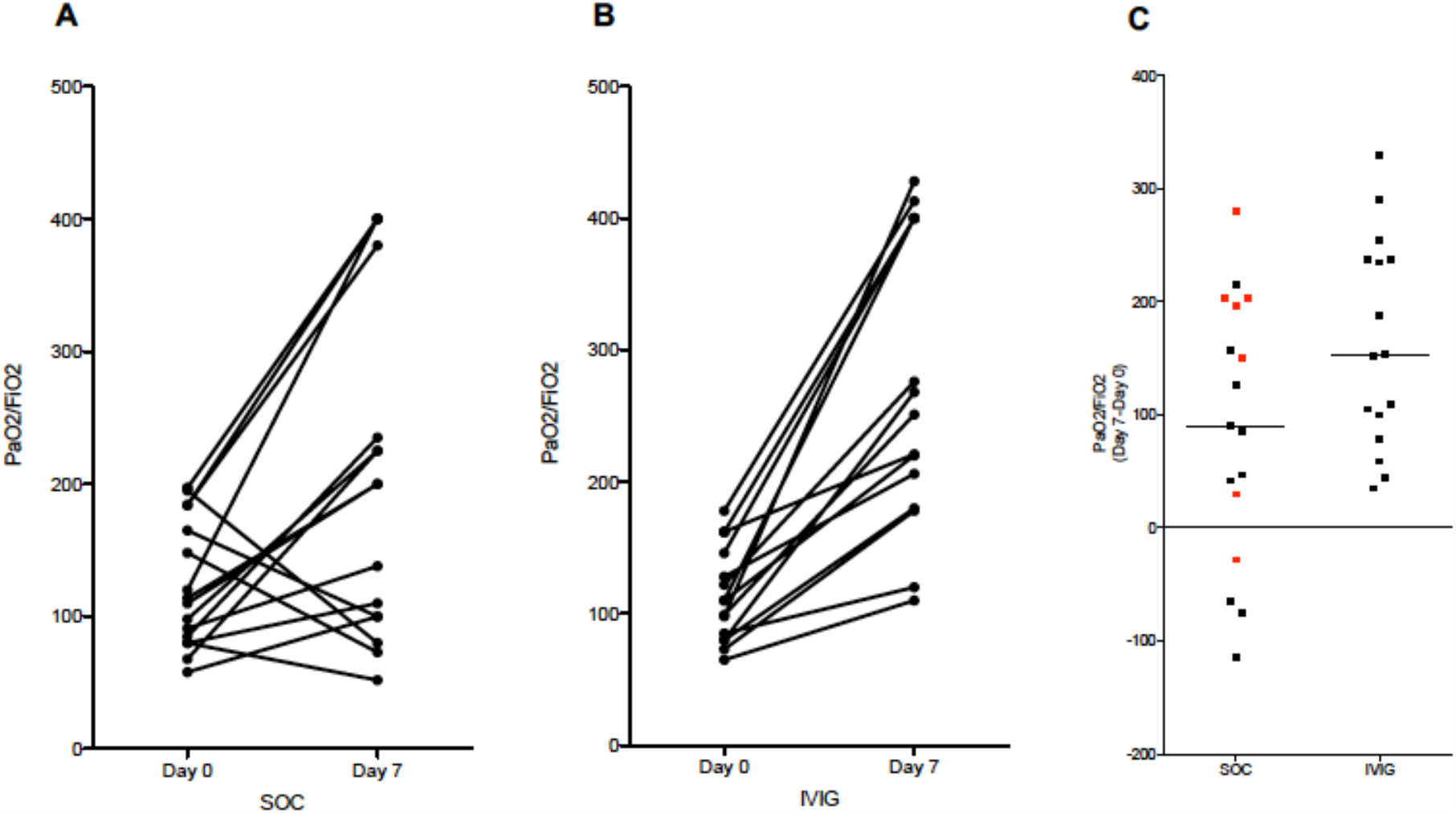
Individual patient progress of PaO_2_/FiO_2_ at day of enrollment (day 0) and 7 days later in control SOC group (A) and IVIG group (B). Patients who were discharged and the one patient that died before day 7 had values placed from the last available day. The absolute differences for each group are shown in part C. Red data points denote patients who did not receive any glucocorticoid therapy. Differences in 7-day PaO_2_/FiO_2_ showed greater improvement oxygenation in IVIG-treated patients when compared to the entire SOC cohort (p=0·057, Mann Whitney U test), but became significant after considering only those patients with A-a gradient at enrollment of >200 (p=0.01, Mann Whitney U) and when comparing IVIG vs only SOC patients who received glucocorticoid therapy (p=0.025, Mann Whitney U).

Figure 4C denotes subjects in the SOC group who did not receive any glucocorticoid therapy as red data points. Seven subjects in SOC did not receive glucocorticoids, 4 of whom were in the low-risk group with A-a gradient < 200. Of the remaining 3 patients, 2 progressed to requiring mechanical ventilation, one of which was made do not intubate and died. PaO_2_/FiO_2_ changes between enrollment and day 7 of the 10 SOC patients who received glucocorticoid therapy (median 11 days) was median +53 (range −115 to +216), a difference that remained significantly lower than the IVIG group (p=0.0057, Mann Whiney U test).

### Adverse events, safety, and tolerability

Three subjects (18%) in the SOC group and one subject (6%) in the IVIG group died. The death in the IVG group occurred after the patient developed a *Staphylococcus aureus* pneumonia and then *Escherichia coli* bacteremia 4 and 6 days after mechanical ventilation, respectively. The SOC deaths included: 1 subject who developed progressive respiratory failure in need of mechanical ventilation who was then made ‘do not intubate’ and expired;; 1 subject had care withdrawn after failing to make progress 20 days on mechanical ventilation;; 1 subject developed *Pseudomonas aeruginosa* and *Enterobacter cloacae* ventilator-associated pneumonia and died after being on mechanical ventilation for 17 days. Tocilizumab was administered off-label to 3 SOC subjects after the endpoint of mechanical ventilation was reached. One subject died, one patient was discharged from the hospital, and one remained in the hospital ventilated at 30 days post-enrollment.

All subjects in the IVIG study arm tolerated IVIG without any adverse events being reported to the clinical study team. Notably, there were no reports of headache, allergic reactions, or thromboses described. All subjects were able to tolerate and complete all 3 daily doses of IVIG without incident. In addition to the bacterial superinfections described above, one additional IVIG subject developed an *E. coli* bacteremia without clear source while otherwise improving clinically from requiring high-flow to low-flow oxygen down to 2 liters by nasal cannula. The patient was prescribed a short course of antibiotics and was discharged home on room air.

### Interleukin-6

A subset of enrolled subjects had serum IL-6 concentrations sent out to a commercial laboratory (ARUP Labs, Salt Lake City, UT) on blood drawn 24-48 hrs post-enrollment. These would be reflective of having received no IVIG in the SOC control arm and 1 or 2 doses of IVIG in the IVIG treatment arm. Figure 5 shows significantly reduced IL-6 serum concentrations in the IVIG group (median 5, range 2-9 pg/mL) compared to the SOC group (median 18, range 3.6-141 pg/mL).

**Figure 5.**
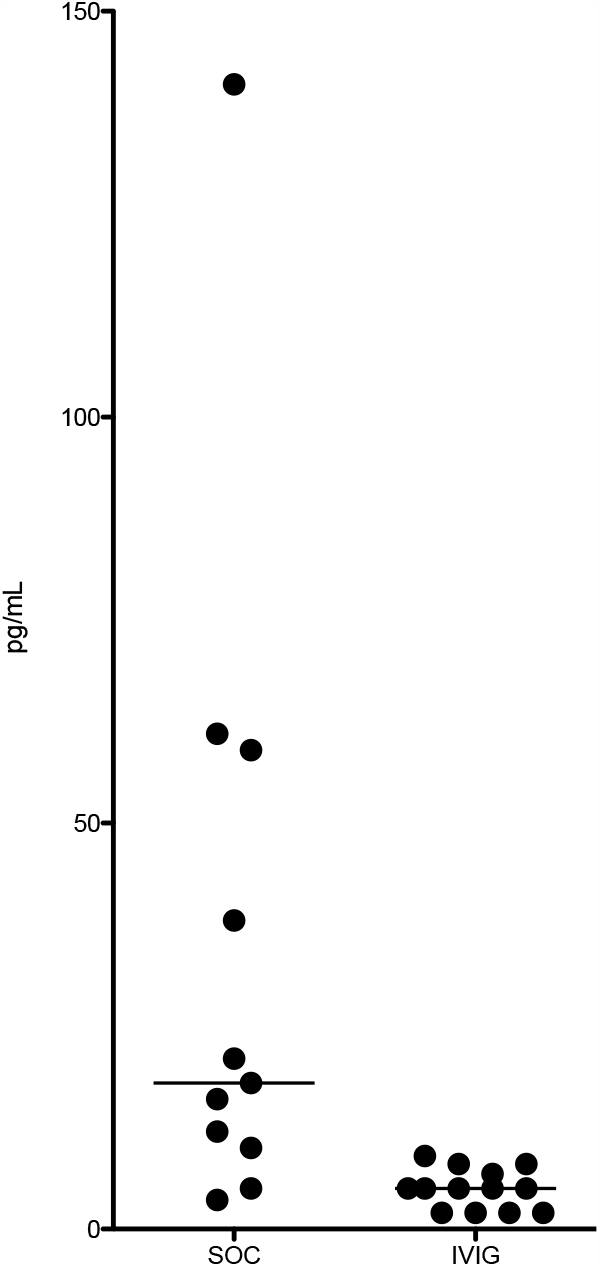
Serum interleukin-6 concentrations of a subset of SOC controls (n=11) and IVIG (n=13) treatment subjects obtained 24-48 hr after enrollment. Median IL-6 18 pg/mL in SOC vs 5 pg/mL in IVIG group are denoted by the horizontal bar (p=0.0012, Mann Whitney u test).

## Discussion

In this first prospective randomized study evaluating IVIG in the treatment of COVID-19 infection, the use of IVIG reduced the rate of progression of respiratory failure requiring mechanical ventilation in COVID-19 patients (13% with IVIG vs. 41% without IVIG, p=0.12). While this did not achieve statistical significance among the collective subject cohorts, the reduced rate of progression to mechanical ventilation with IVIG achieved statistical significance among the subset with a calculated or estimated A-a gradient of > 200 mm Hg (14% with IVIG vs. 58% without IVIG, p=0.038). This finding points not only to the potential benefit of IVIG in COVID-19, but the importance of optimal patient selection for IVIG in maximizing clinical benefit. COVID-19 patients with mild hypoxia may never warrant anything more than supportive low-flow oxygen therapy, such that intervening with IVIG may not change the course of an otherwise benign disease course. A cutoff A-a gradient of >200 mm Hg was chosen due to the fact that this represents a notable risk increase in APACHE 2 score calculation in predicting mortality. The benefits of IVIG in reducing respiratory failure morbidity in COVID-19 patients was further supported by the significant shortening of duration of length hospital stay and ICU length of stay, and improvement in oxygenation (PaO_2_/FiO_2_) at day 7. Mortality was numerically reduced from 3/17 (18%) to 1/16 (6%) with IVIG in this study but not significant due to small sample size.

A multisystem inflammatory syndrome (MIS) has been recently described in children and adolescents convalescing from COVID-19 infection. This syndrome which includes fever, rash, myocardial dysfunction, gastrointestinal disturbance, hypotension, coagulopathy, and elevated blood markers of inflammation (eg. C-reactive protein) is clinically reminiscent of Kawasaki’s disease, a well-recognized idiopathic severe febrile illness of childhood.^23,24^ This raises the possibility that the pathophysiology of COVID-19 infection in children and adults may be similar even though clinical manifestations are different. Interestingly, the mainstay of therapy for Kawasaki disease for decades has been IVIG, and children with MIS have been successfully treated with IVIG, lending further support of a potential benefit of IVIG in adults with COVID-19.^17,23,24^

Although the sample size was small, IVIG was well-tolerated in this study and did not require discontinuation and the three-day course was completed by all subjects. The hypercoaguability conferred by IVIG raised theoretical concerns when superimposed on the recently observed COVID-19 induced thrombotic events, but no cases of arterial or venous thrombosis associated with IVIG were identified in this study. This is particularly notable given that 15 of the 16 subjects in the IVIG treatment arm had elevated D-dimer concentrations at the time of enrollment, suggestive of some form of intravascular thrombosis and fibrinolysis consistent with the pathophysiology of more severe COVID-19 cases. Furthermore, antiphospholipid antibodies have been described in COVID-19 patients and IVIG has been used has been used previously to treat the antiphospholipid antibody syndrome.^**25**,**26**^

The importance of IL-6 in the cytokine release syndrome (CRS) in COVID-19 has been suggested by the Th17 lymphocyte populations found in patients with alveolar injury, initiating the study of IL-6 receptor blockade in the treatment of severe COVID-19 infection.^11-14,27^ Data appears promising but randomized control data beyond descriptive case series is still awaited. In this study, the subset of patients who had IL-6 concentrations measured within 24-48 hours of study enrollment showed significantly lower IL-6 levels among those who received IVIG compared to the SOC group. While pre-enrollment IL-6 levels were not measured, reduction of IL-6 production by IVIG has been previously shown and further validates the potential role of IVIG in severe COVID-19 infection.^28^

As a blood product derived from healthy donors, efficacy of IVIG may in fact improve over time as a higher percentage of the population of donors develops neutralizing antibodies to the SARS-CoV2 virus. Convalescent plasma may offer some benefit in a subset of COVID-19 patients per a recent randomized study, so these benefits could theoretically be added to the immunomodulatory effects of IVIG as the donor pool of IVIG develops increased immunity to SARS-CoV-2.^29^

This study has some important limitations. First, this study was performed in just 2 hospitals in one US city, resulting in a fairly homogenous population of younger Latino patients where results may not automatically translate to other patient settings. Secondly, while prospective and randomized, the study was not blinded and therefore subject to bias. This was reduced by the fact that the clinical investigators had minimal clinical decision-making on these patients, particularly regarding the endpoints of mechanical ventilation and discharge from the hospital. Third, the concomitant use of methylprednisolone therapy may have confounded the results. At the time of study initiation, the use of glucocorticoid therapy was controversial, possibly causing harm in COVID-19. We decided to give methylprednisolone 40 mg (equivalent to approximately 7·5 mg dexamethasone) to mitigate any potential adverse effects of IVIG such as headache, theorizing that benefit would outweigh risk by increasing tolerability of IVIG. Just recently, however, the benefit of glucocorticoid therapy was shown with 6 mg of dexamethasone for 10 days.^30^ While there may have been a small benefit rendered by this premedication, it is unlikely that the difference in outcomes between the two groups rested on this intervention. This is supported by the fact that the benefits of IVIG were significant even when comparison was made to the subset of SOC patients that received glucocorticoid therapy. Fourth, the sample size was small, which markedly reduced the power and strength of the positive findings, particularly given that COVID-19 treatment standards have been a moving target. The 6-week study enrollment period saw a shifting attitude towards favoring glucocorticoids, the availability of remdesivir, and the movement away from early intubation/mechanical ventilation in favor of more aggressive self-proning protocols.^31,32^ Finally, severe comorbidities like severe renal disease and heart failure that were not highly represented in this group of patients may pose particular challenges due to their potentially poor tolerability of the volume load of IVIG therapy during COVID-19 infection.

In summary, this pilot study showed that IVIG 0.5g/kg daily for 3 days reduced progression of respiratory failure requiring mechanical ventilation, total length of hospital stay, and ICU length of stay, and improved oxygenation at 7 days when given after premedication with methylprednisolone 40 mg to COVID-19 patients with a calculated or estimated A-a gradient of > 200 mm Hg (PaO_2_/FiO_2_ <140, PaO_2_ ≤ 92% on ≥ 6 L nasal cannula). This study served as the foundation of a larger phase 3, multicenter, double-blind placebo-controlled trial evaluating IVIG in COVID-19, which is currently enrolling patients (clinicaltrials.gov, NCT04400058), and which will hopefully validate these findings.

## Data Availability

Data is currently under peer review. Crude data is available without patient identifying information..

## Acknowledgments

Guarantor statement: George Sakoulas, MD takes responsibility for (is the guarantor of) the content of the manuscript, including the data and analysis

## Author Contributions

Clinical trial design: GS, MG, FH Clinical trial execution: GS, MG, FH

Data analysis: GS, MG, RK, KLG, MH, AV, MG(2), VNKD, FH

Manuscript First Draft: GS

Manuscript Editing: GS, MG, RK, KLG, MH, AV, MG(2), VNKD, FH

Figures: GS

## Financial Disclosures

This research was supported and funded by Octapharma USA, Inc (Paramus, NJ).

Octapharma USA, Inc did not influence the design or execution of the study, and did not have a role in data analysis or manuscript preparation.

---

**Supplemental Figure.**
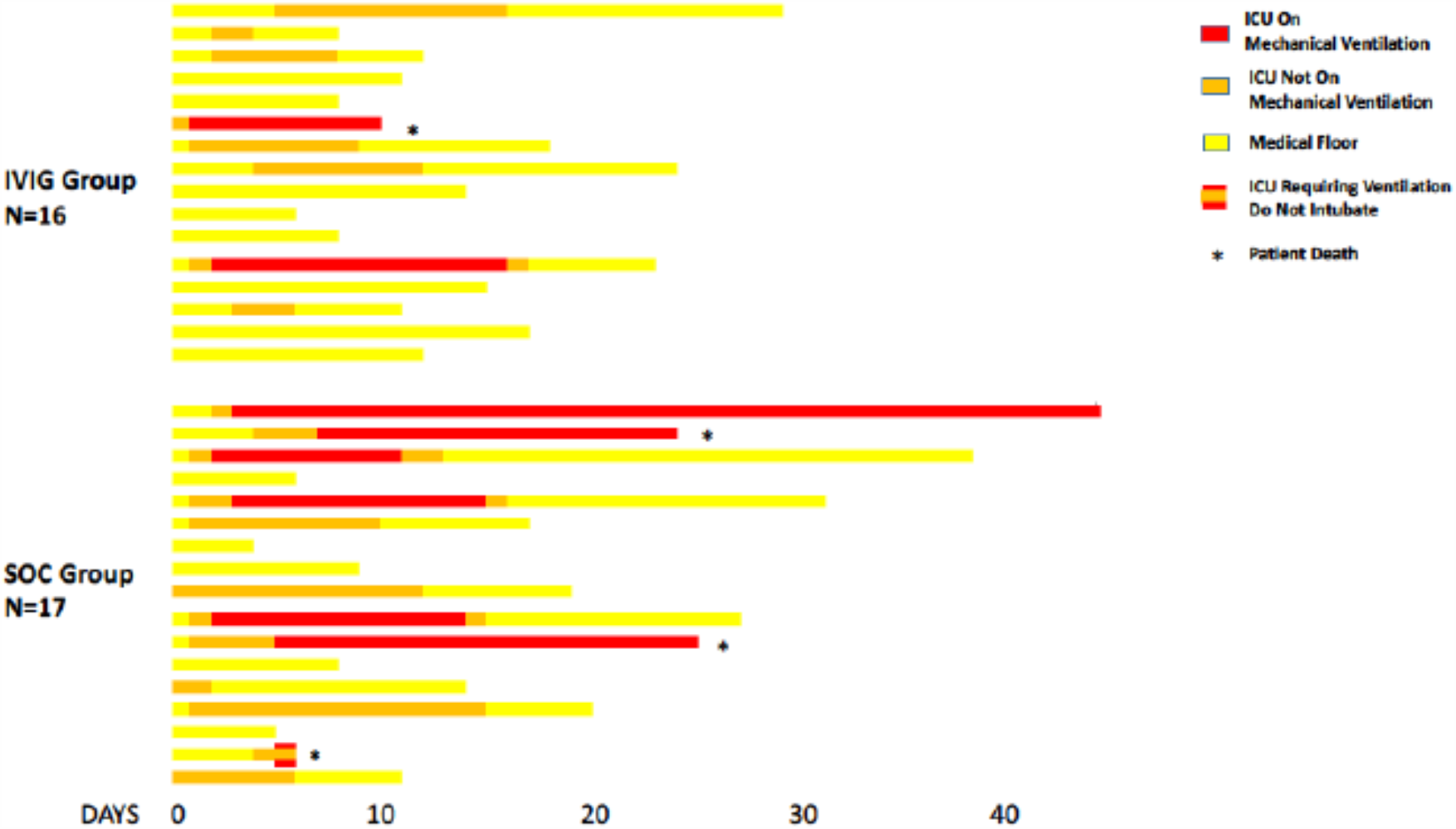
Schematic diagram of hospitalization from the time of admission of subjects. Death occurred in 3 SOC subjects and 1 IVIG subject (*).

